# Multi-center Clinical Study Using Optical Coherence Tomography for Evaluation of Cervical Lesions In-vivo

**DOI:** 10.1101/2020.05.12.20098830

**Authors:** Chenchen Ren, Xianxu Zeng, Zhongna Shi, Chunyan Wang, Huifen Wang, Xiuqin Wang, Baoping Zhang, Zhaoning Jiang, Huan Ma, Hao Hu, Haozhe Piao, Xiaoan Zhang, Baojin Wang

## Abstract

**Objective:** In this prospective study of an in-vivo cervical examination using optical coherence tomography (OCT), we evaluated the diagnostic value of non-invasive and real-time OCT in cervical precancerous lesions and cancer diagnosis, and determined the characteristics of OCT images.

**Methods:** 733 patients from 5 Chinese hospitals were inspected with OCT and colposcopy-directed biopsy. The OCT images were compared with the histological sections to find out the characteristics of various categories of lesions. The OCT images were also interpreted by 3 investigators to make a 2-class classification, and the results were compared against the pathological results.

**Results:** Various structures of the cervical tissue were clearly observed in OCT images, which matched well with the corresponding histological sections. The OCT diagnosis results delivered a sensitivity of 87.0% (95% confidence interval, CI, 82.2%-90.7%), a specificity of 84.1% (95% CI, 80.3%-87.2%), and an overall accuracy of 85.1%.

**Conclusion:** Both good consistency of OCT images and histological images and satisfactory diagnosis results were provided by OCT. Due to its features of non-invasion, real-time, and accuracy, OCT is valuable for the in-vivo evaluation of cervical lesions and has the potential to be one of the routine cervical diagnosis methods.

## Introduction

Cervical cancer is one of the most common malignant tumors of the reproductive system in women. It is one of the main gynecological tumors causing female death with a low 5-year survival rate [1]. Clinical reports show that the global incidence and mortality rate of cervical cancer is 6.6% and 7.5%, respectively, while 85% of cervical cancer cases are in developing countries [2]. At present, the incidence of cervical cancer shows a trend of getting younger. Studies show that the development of cervical cancer needs a relatively long time from several years to decades, including the vital stage of cervical intraepithelial neoplasia (CIN). So early screening and treatment of precancerous lesions play important roles in preventing cervical cancer [3]. The wide application of effective cervical cancer screening in developed countries has led to a significant reduction in cervical cancer incidence [4].

Currently, there are several clinical screening and diagnosis methods to identify patients with precancerous lesions, including Pap smear, human papillomavirus (HPV-DNA) test, ThinPrep liquid-based cytology test (TCT), colposcopy, and cervical biopsy. As the earliest cytological test method, Pap smear has been gradually replaced by TCT due to its low accuracy, although it costs much less [5]. HPV tests have been widely used in clinical practice. HPV test has a high sensitivity but a low specificity, which leads to a fairly high rate of false positives [6, 7]. High sensitivity means that HPV tests can reduce the occurrence of missed diagnosis, but too many misdiagnoses occur because HPV infections can be transient and may not develop to cervical epithelial lesions in most cases. TCT is also widely used in clinical practice as a cervical cytology testing technique, which has a higher specificity than the HPV test. However, it has a much lower sensitivity for CIN detection compared with the HPV test [8]. Factors such as inflammation, unsatisfactory sampling, and lack of experience of a cytologist may all lead TCT to a missed diagnosis.

Colposcopy is a visual inspection method utilizing the magnification function of a colposcope. It gives a direct-viewing impression of the cervix, but its accuracy relies heavily on the experience of the investigators. A study shows that the sensitivity of conventional colposcopy was merely 55% [9]. Colposcopy-directed biopsy with histopathological confirmation is the current gold standard for diagnosing cervical diseases, but biopsy is invasive and may cause bleeding and infection. Moreover, since only a few cervical locations can be selected for biopsy, the chance of missed diagnoses of highrisk lesions is as high as 37% [10].

Optical coherence tomography (OCT) is a 3-D imaging technology first developed in the 1990s [11]. Based on the principle of low coherent light interference, the imaging of superficial biological tissue is carried out by detecting the interference signal formed by a reference light and back-scattered light from different depths of the sample. After the back-scattered light signal is detected and processed, 3-D images of the internal tissue microstructures can be obtained. The axial resolution of ~1-10 μm can be generally reached, while the transverse resolution can also reach a few microns [12]. OCT can detect tissue structures with a penetration depth of 1-2 mm [13]. Although the imaging depth is relatively shallow compared to other clinical imaging methods, such as ultrasound, MRI, and CT, the resolution of OCT in tissues is 50-100 times higher than that of ultrasound, far higher than the resolution of MRI and CT. With its advantages of high resolution, rapid imaging, and non-invasiveness, OCT has attracted much attention in clinical applications. Currently, it has been widely applied to examinations of ophthalmic [14], cardiovascular [15], and oral diseases [16].

In a previous study by Zeng et al., an optical coherence microscopy (OCM) system was used to analyze the image features of cervical tissue and it demonstrated satisfactory diagnosis results in a 2-class classification analysis with ex-vivo cervical tissues [17]. In this study, we performed a multi-center clinical study to obtain OCT images of in-vivo cervical tissues. By comparing the OCT images with corresponding histological images, the optical features of various categories of in-vivo cervical lesions were summarized. Then, the OCT diagnosis results were analyzed, and the clinical application value of OCT on the diagnosis of in-vivo cervical lesions was discussed.

## Methods

### Clinical OCT System

The OCT system adopted in this study is Ultralucia OCT Cervical Scanning System (model: UL-C100) developed by Zhengzhou Ultralucia Medical Technology Co., Ltd. This device is equipped with a broadband light source with a central wavelength of 850 nm, providing an axial resolution of <5 μm and a transverse resolution of <10 μm in tissue. A handheld probe (shown in Figure 1) was used to deliver the imaging beam to the surface of the cervix. The outer diameter of the probe is ~10 mm, making it easy to pass through the speculum. The maximum scanning speed is 80,000 A-scans/s, and the imaging depth is ~1 mm. In our study, a circular scanning mode is adopted for image collection, and the scanning diameter can be adjusted within the range of 0 to 2 mm according to the setting. Compared to the traditional linear scanning mode, the circular scanning mode can obtain a much wider scanning range with the thin imaging probe, allowing more tissue information to be collected in each frame of the OCT image. In this study, the number of A-scans per frame is set to 1200. The scanning diameter is 0.9 mm (the inner circle) to 1.1 mm (the outer circle).

**Figure 1.**
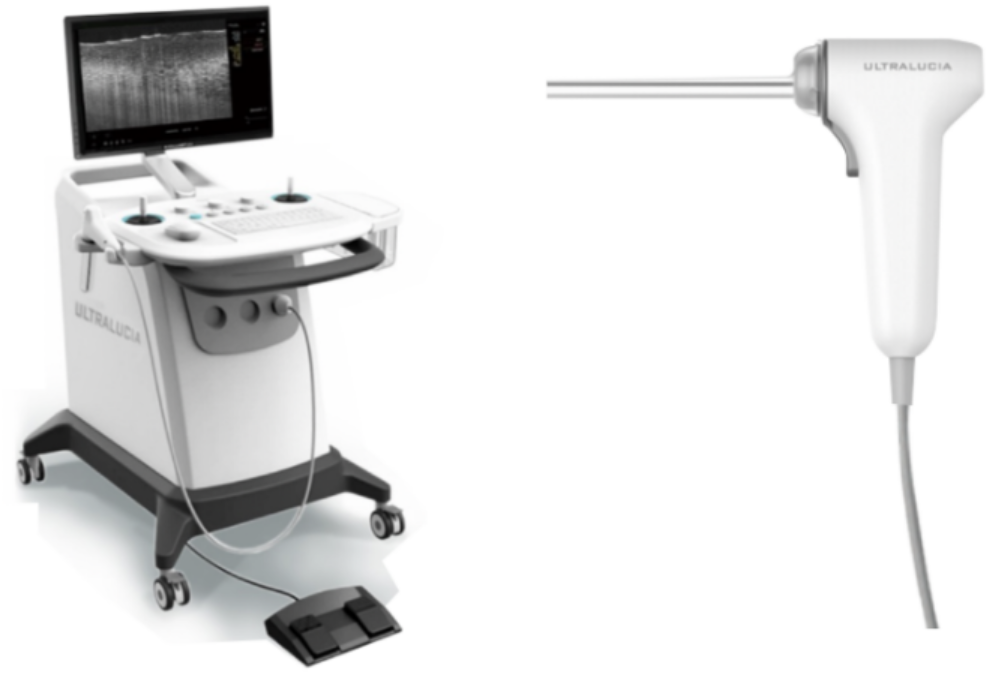
Ultralucia OCT Cervical Scanning System and Handheld Probe.

### Study Plan and Examination Process

The study was approved by the Ethics Committee of the Third Affiliated Hospital of Zhengzhou University, the Ethics Committee of Liaoning Cancer Hospital & Institute, the Ethics Committee of Puyang Oilfield General Hospital, the Ethics Committee of Luohe Central Hospital, and the Ethics Committee of Zhengzhou Jinshui District General Hospital. All research was performed in accordance with relevant guidelines/regulations. Informed consent was obtained from all participants and/or their legal guardians.

733 gynecological outpatients were recuited in the above five Chinese hospitals from Aug 2017 to Dec 2019, all of which had signed informed consent. These hospitals consist of two provincial hospitals, two municipal hospitals, and one district hospital. All the recruited patients had either positive HPV results or suspicious cytology findings of ASCUS (atypical squamous cells of undetermined significance) or above, or both. Then, the patients were inspected with in-vivo OCT and received colposcopy-directed cervical biopsy. Table 1 shows demographic information of the patients.

**Table 1.** Patient Demographic Information

| Hospital | No. of Patients | Age (mean ± SD) | HPV Results | TCT Results |
| --- | --- | --- | --- | --- |
| The Third Affiliated Hospital of Zhengzhou University | 350 | 38.67±9.86 | Positive: 281<br>Negative: 31<br>Untested: 38 | Positive: 215<br>Negative: 63<br>Untested: 72 |
| Liaoning Cancer Hospital & Institute | 227 | 44.08±8.38 | Positive: 138<br>Negative: 84<br>Untested: 12 | Positive: 69<br>Negative: 134<br>Untested: 24 |
| Puyang Oilfield General Hospital | 59 | 43.03±8.06 | Positive: 39<br>Negative: 3<br>Untested: 17 | Positive: 39<br>Negative: 12<br>Untested: 8 |
| Luohe Central Hospital | 57 | 40.37±10.05 | Positive: 49<br>Negative: 8<br>Untested: 0 | Positive: 36<br>Negative: 21<br>Untested: 0 |
| Zhengzhou Jinshui District General Hospital | 40 | 39.03±12.47 | Positive: 36<br>Negative: 2<br>Untested: 2 | Positive: 9<br>Negative: 27<br>Untested: 4 |
| Overall | 733 | 40.85±9.79 | Positive: 536<br>Negative: 128<br>Untested: 69 | Positive: 368<br>Negative: 257<br>Untested: 108 |

After consent, the examination process is as follows: (1) Preparation: Insert a speculum to the vagina of the patient. Fully expose the cervix, then wipe off any secretion, and make a preliminary visual observation. (2) OCT scanning: Mount a disposable protective cover to the handheld probe of the OCT system. Gently contact the cervical surface with the probe, then scan 12 locations in clockwise order and save the OCT images. (3) Colposcopy inspection: Firstly observe the whole picture of the vagina and cervix with a low-power lens, and observe the cervix with a high-power lens. Then, apply glacial acetic acid and compound iodine solution in turn, observe the cervix focusing on the squamocolumnar junction (SCJ) and the transformation zone. (4) Biopsy: Take biopsies at the suspicious locations based on the colposcopy impression, or take biopsies at 3, 6, 9, and 12 o’clock locations of the cervix in atypical cases. Finally, send the biopsy samples for pathological examination.

### Image Processing and Comparison

To compare OCT images with corresponding histological images, firstly, we scanned the H&E sections with a MoticEasyScan scanner (model: BA600Mot) and obtained the digital histological images of all the biopsy locations. Then, the digital H&E images were magnified at 10x magnification for comparison. Meanwhile, we processed OCT images of the corresponding locations. In PowerPoint, the OCT images were appropriately scaled and adjusted to a ratio of 1:1 in the axial and lateral directions. For each biopsy location, the OCT image and the corresponding histological image were placed side by side in PowerPoint on the same scale. By comparing the images carefully, we acquired many OCT-H&E image pairs with a good match and discovered the OCT optical features of various lesion categories.

### OCT Image Classification and Statistical Analysis

The OCT image feature interpretation used in this study was adapted from the classification criteria described previously [17]. For every scanning location, a 2-class classification was made according to the observed features from OCT images. We defined normal/mild inflammation, ectropion, and low-grade squamous intraepithelial lesion (LSIL) as low-risk or negative, and high-grade squamous intraepithelial lesions (HSIL) and invasive lesions as high-risk or positive. Three trained investigators used ImageJ image processing software (version 1.51j8) to review the OCT images without knowing the corresponding histological section and diagnosis information. Only patient information of the age and HPV/TCT results were provided to the investigators. For each patient, a positive result was given if at least one location was marked as high-risk. Otherwise, a negative result was given. The third investigator was responsible for the result check. In case opposite results were given by the first two investigators, the third investigator made the final decision. H&E sections of the biopsy samples were prepared and reviewed by local pathologists in each hospital. The sensitivity, specificity, and accuracy of OCT were calculated by comparing OCT classification results against the pathological results.

## Results

### Diagnostic Features of OCT Images

Diagnostic features of OCT images are identified in OCT images in comparison with corresponding histological sections. Based on the optical features, low-risk and high-risk cervical lesions can be distinguished. Figure 2 shows OCT images and corresponding H&E histological sections of low-risk tissues, which includes normal/mild inflammation, ectropion, and LSIL. Since a disposable protective cover was applied to the handheld probe when scanning in-vivo tissues, a thin layer of the cover, as well as the outer surface of the probe window, may be observed on the top of cervical tissue in OCT images.

Figure 2A-C display a case of normal cervical tissue. Figure 2A is the colposcopy image, and the small green circle shows the location where Figure 2B-C were acquired. In this case, the HPV result is positive, and the TCT result is ASCUS. While the OCT image interpretation delivered negative, which is consistent with the pathology result. As for normal cervical tissues, the OCT images are uniform in refraction from top to bottom. Squamous epithelium (EP) cells are arranged in a well-organized way forming a mesh-like structure. Squamous epithelium and stroma (ST) are well stratified so that the whole basal membrane (BM) between epithelium and stroma can be observed clearly in OCT images. Glandular structures can be identified in the stroma as strip-type hypo-scattering regions.

**Figure 2.**
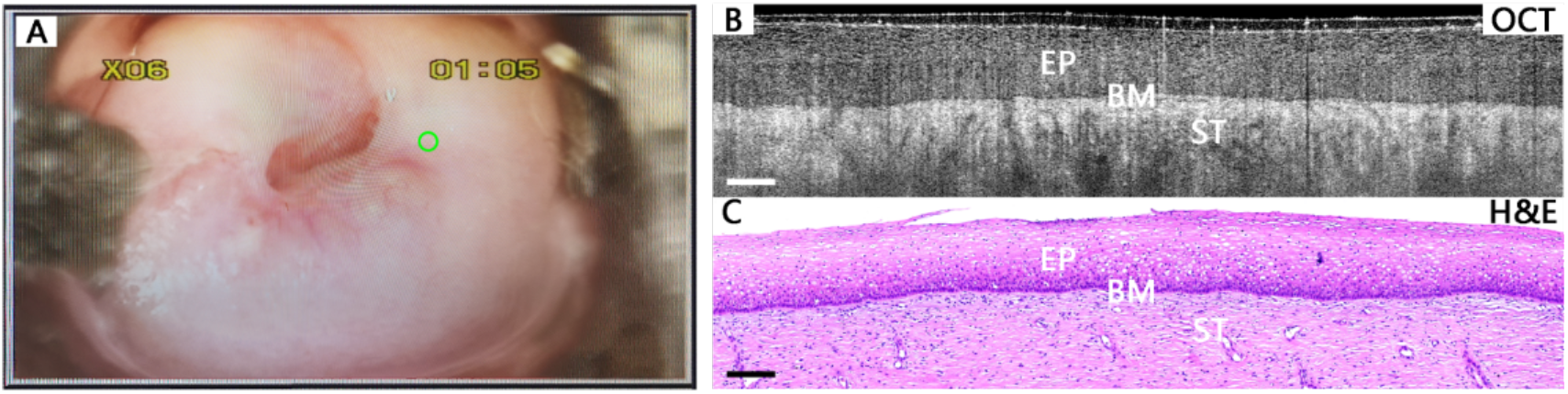
A typical case of normal cervical tissue. Colposcopy image (A), OCT image (B), and the corresponding histological image (C). The basal membrane (BM) can be clearly visualized, which forms the boundary between the epithelium (EP) and stroma (ST). Scale bars: 200 μm. H&E magnification: 10x.

Figure 3 shows more cases of low-risk cervical tissues. In some inflammation tissues as shown in Figure 3A-B, the layered structure is similar to normal cases, but Nabothian cysts (NC) can be identified as large hypo-scattering regions under the stroma layer. Figures 3C-D present OCT images and corresponding H&E histological sections of cervical columnar epithelial ectropion. Stratified squamous epithelium structure can no more be observed in OCT images. Instead, columnar epithelium cells form regular papillary or glandular structures in ectropion tissues. The typical structural feature can also be visualized in H&E histological sections. Though epithelial LSIL tissues (Figures 3E-F) show epithelial structure very similar to normal cervical tissues, some koilocytotic cells can be identified in OCT images. Enlarged cell nuclei can be observed as hyper-scattering spots, while enlarged perinuclear halos result in big hypo-scattering cells in OCT images. The appearance of this structure feature shows HPV infection in the cervical tissue.

**Figure 3.**
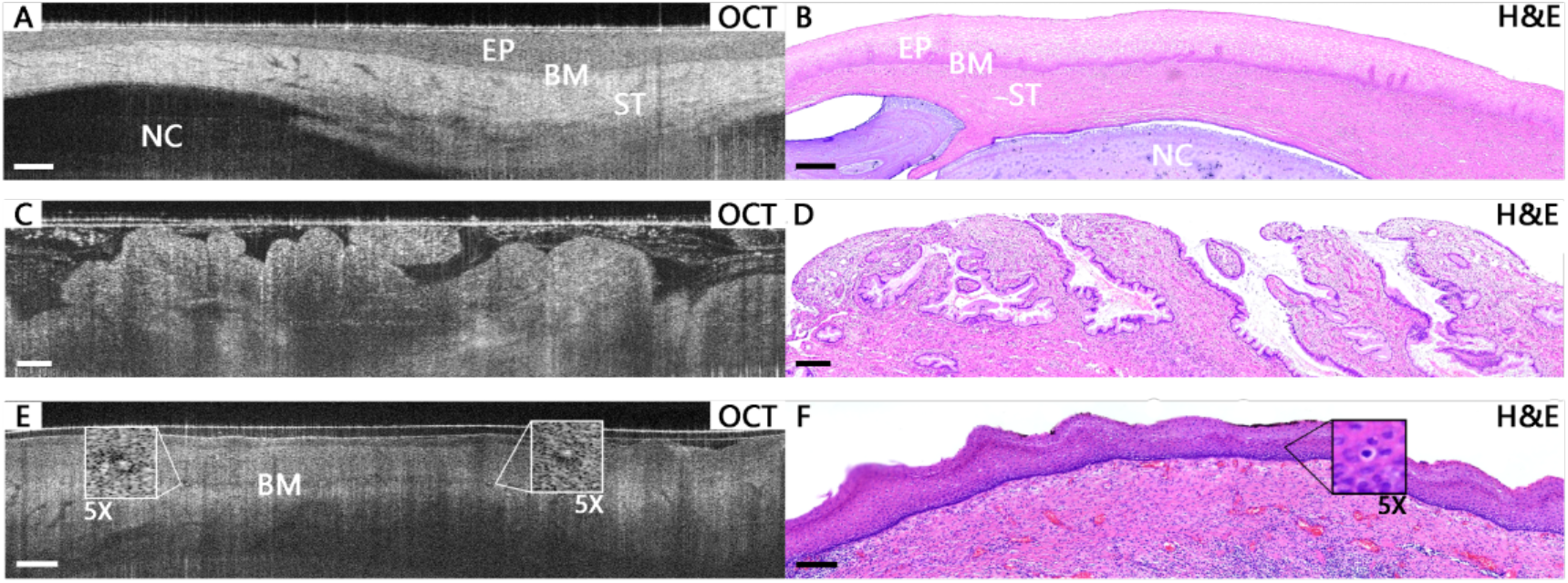
Typical OCT images and corresponding H&E histological images of low-risk cervical tissues. In (A) and (B), a Nabothian cyst of inflammation tissue can be observed under the stroma layer. In ectropion tissue, the papillary structure with hyper-scattering boundaries is visible in the OCT image (C), which is consistent with the single-layered columnar cells in the H&E section (D). In an LSIL tissue, koilocytotic cells are clearly visible at additional 5x magnification in OCT image (E) and H&E section (F). Scale bars: 200 μm. H&E magnification: 10x.

Figure 4 shows well-matched OCT images and corresponding H&E histological sections of HSIL cervical tissues. Figure 4A-C present an HSIL case, which has a positive HPV, and the TCT result shows HSIL not excluding invasive cancer. In the colposcopy image (Figure 4A), the small green circle shows the OCT scanning location where Figure 4B-C were acquired. Since more than one-third of the squamous epithelium cells in the epithelial layer is affected, the orderly epithelial structure can no longer be observed. In this case, as shown in Figure 4B, the layered structure and the basal membrane are completely invisible. The image intensity is high at the surface and decreases rapidly with the increase of depth. Hyper-scattering icicle-like features (arrows) can usually be observed at the bottom of the bright layer. In some other cases, as shown in Figure 4D, the epithelium structure is disorganized with several hypo-scattering regions in the epithelial layer, but the basal membrane is still partially visible.

**Figure 4.**
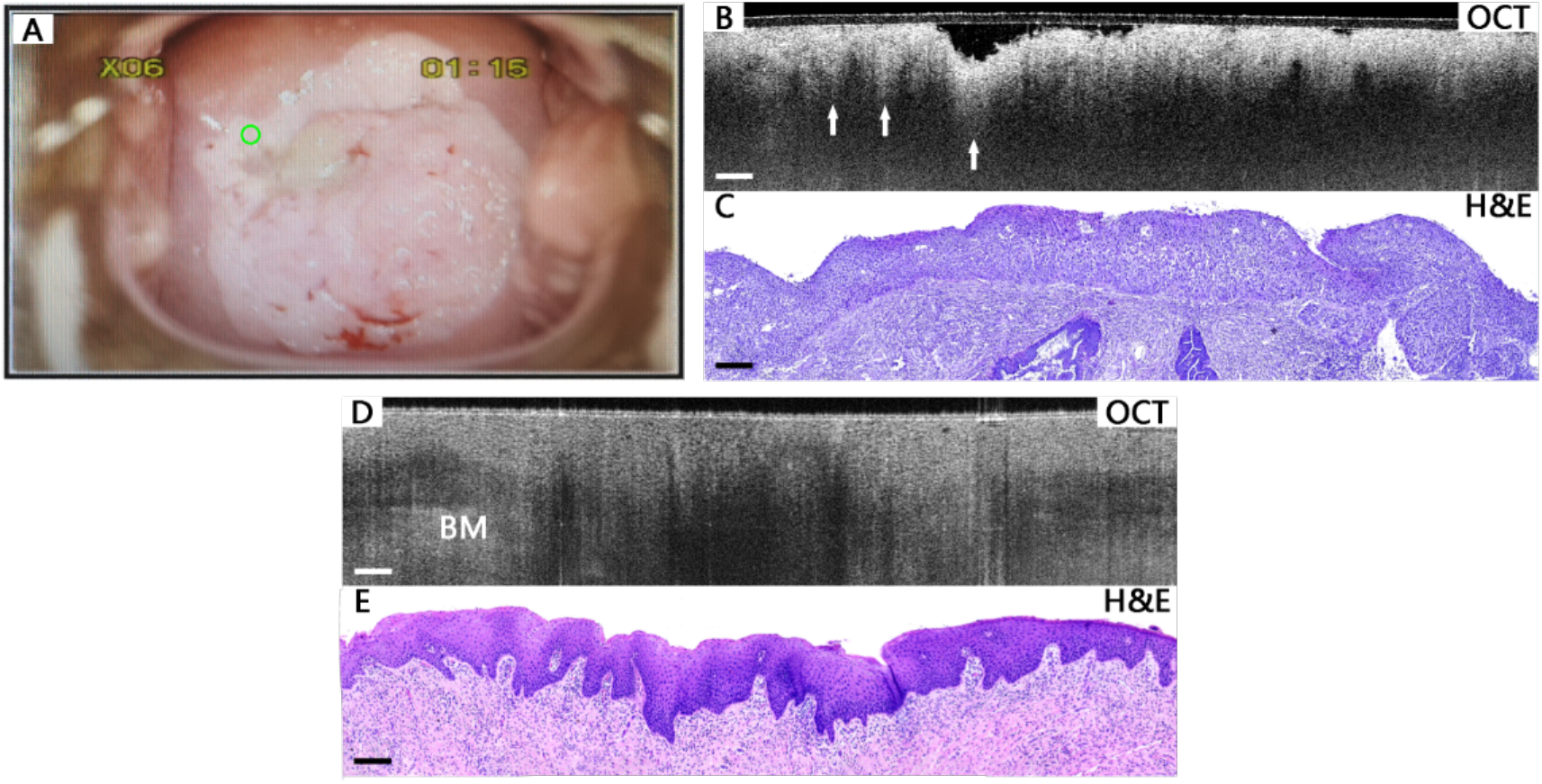
Typical images of HSIL cervical tissues. Colposcopy image (A), OCT image (B), and the corresponding histological image (C) of an HSIL case. In the OCT image (B), the layered structure is lost, and the intensity is rapidly attenuated downwards, with hyper-scattering icicle-like feature (arrows) visible. OCT images (D) and corresponding H&E histological sections (E) of another HSIL tissue. In the OCT image (D), the layered structure still exists, hypo-scattering regions can be observed in the epithelial layer, and the basal membrane can still be found but less clear. Scale bars: 200 μm. H&E magnification: 10x.

Figure 5 shows image comparisons of cervical invasive lesion tissues. Figure 5A-C present a cervical squamous carcinoma case with positive HPV result. Similar to HSIL, invasive lesions also lose the layered structure in the OCT images. Neither epithelium nor basal membrane can be observed in OCT images. Heterogeneous regions of hypo-scattering (Figure 5B) or hyper-scattering nests or clusters of squamous cell tumors can be identified as typical features of cervical invasive lesions. In some invasive lesion cases (Figure 5D), the OCT images are very similar to the HSIL case shown in Figure 4B, with high-intensity surface layer and icicle-like features. Table 2 shows a summary of the optical features of various cervical lesions.

**Figure 5.**
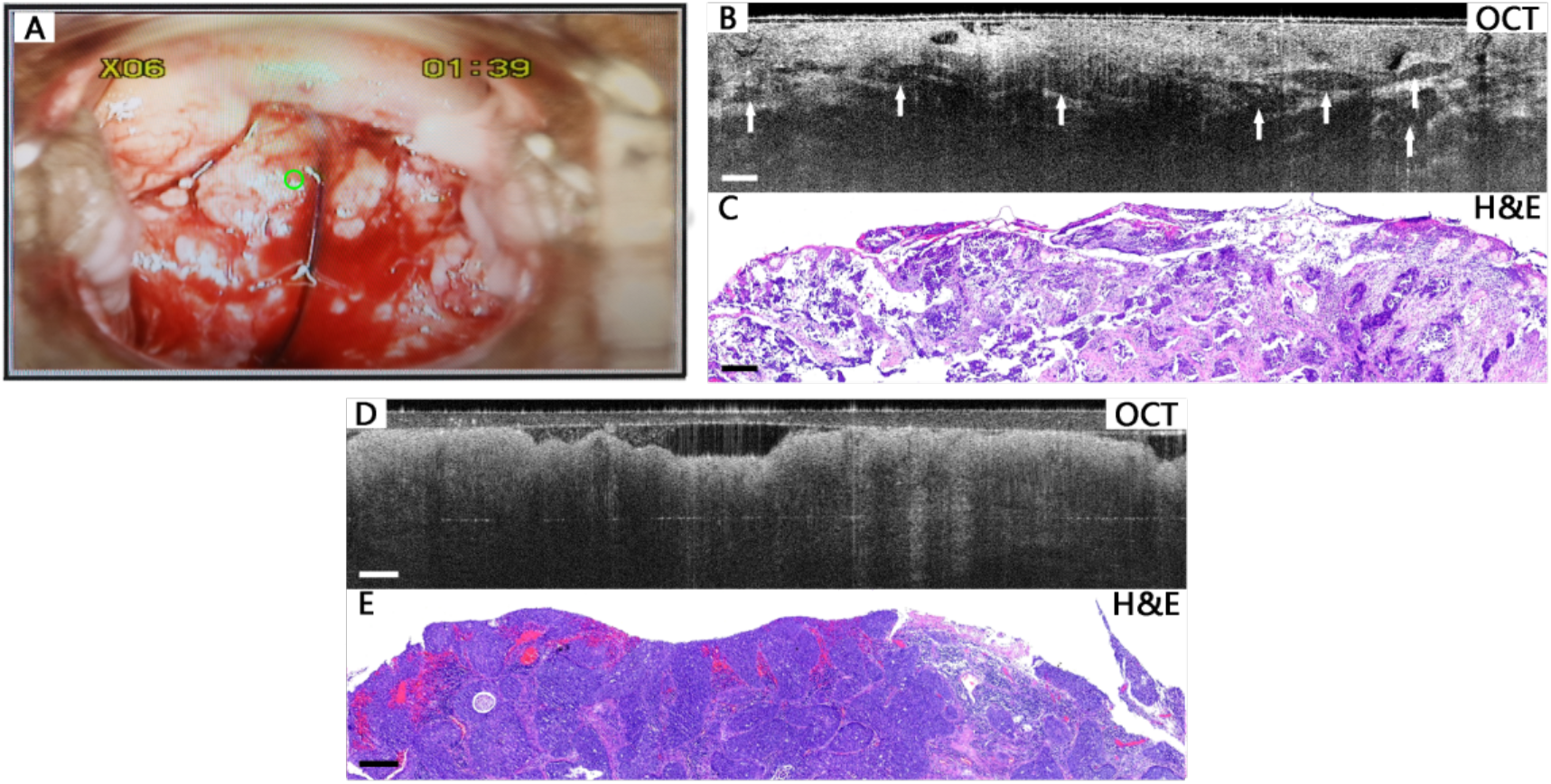
Typical images of cervical cancer tissues. Colposcopy image (A), OCT image (B), and the corresponding histological image (C) of a squamous carcinoma case. In the OCT image (B), heterogeneous regions of hypo-scattering nests or clusters (arrows) can be identified. OCT images (D) and corresponding H&E histological sections (E) of another cancer tissue. In the OCT image (D), similar to Figure 4B, hyper-scattering icicle-like feature is visible. Scale bars: 200 μm. H&E magnification: 10x.

**Table 2.**
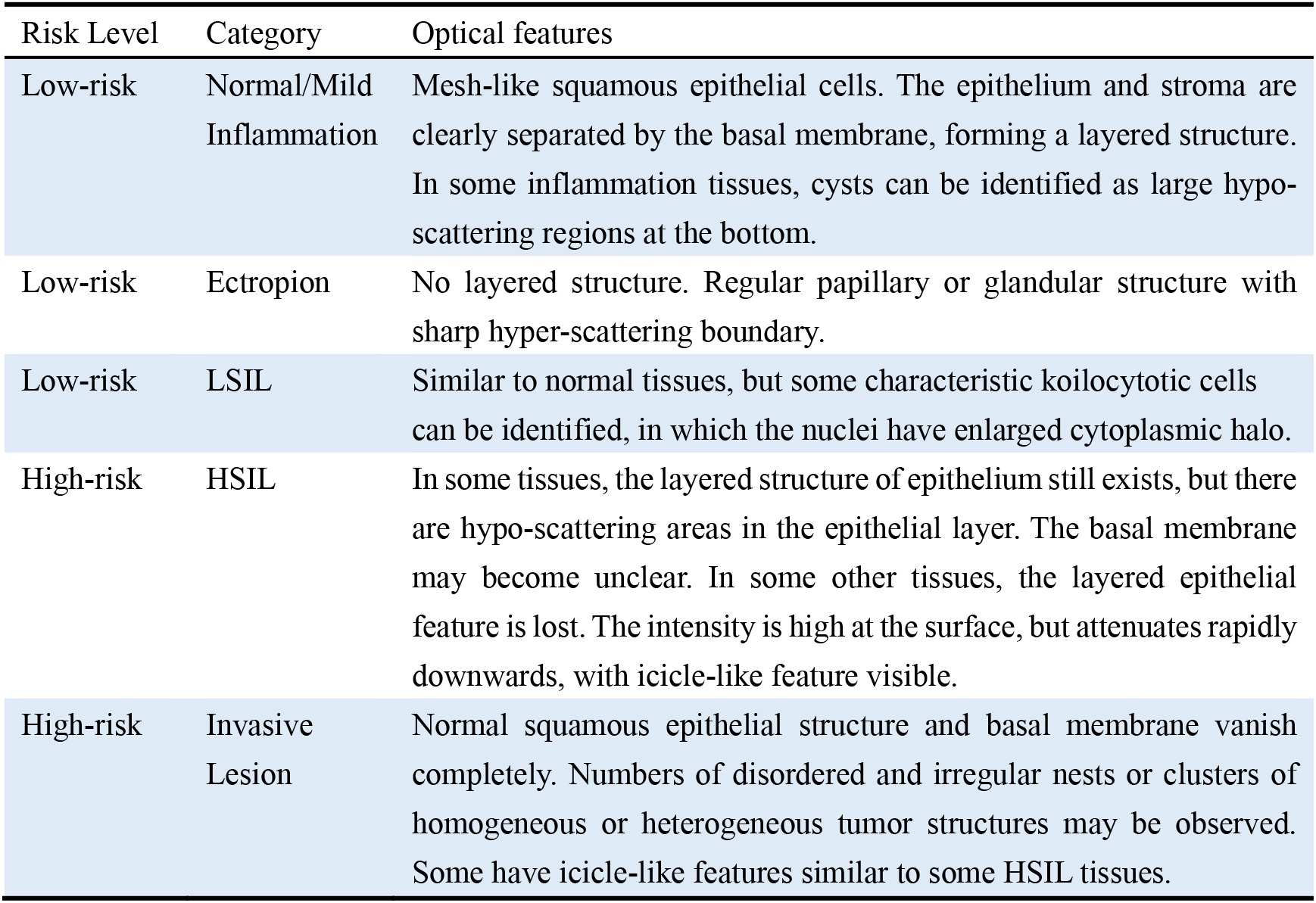
Optical features of different cervical lesions on OCT

### Image Classification and Statistics Analysis

Among the 733 patients recruited in this study, the OCT test results of 624 patients are consistent with the pathological results, among which 234 have positive results, and 390 are negative. The overall OCT diagnosis turned out a sensitivity of 87.0% (95% CI, 82.2%-90.7%), a specificity of 84.1% (95% CI, 80.3%-87.2%), and an overall accuracy of 85.1%. The values of 95% CI were calculated according to the efficient-score method described by Robert Newcombe [18], based on the procedure outlined by E. B. Wilson [19] in 1927. Table 3 summaries the diagnosis results of all the hospitals.

**Table 3.** OCT Diagnosis Results

| Hospital | No. of Patients | Sensitivity (95% CI) | Specificity (95% CI) | Accuracy |
| --- | --- | --- | --- | --- |
| The Third Affiliated Hospital of Zhengzhou University | 350 | 84.9%<br>(77.2%-90.4%) | 85.7%<br>(80.3%-90.0%) | 85.4% |
| Liaoning Cancer Hospital & Institute | 227 | 87.8%<br>(78.8%-93.4%) | 76.6%<br>(68.5%-83.3%) | 81.1% |
| Puyang Oilfield General Hospital | 59 | 85.7%<br>(66.4%-95.3%) | 87.1%<br>(69.2%-95.8%) | 86.4% |
| Luohe Central Hospital | 57 | 100.0%<br>(79.1%-100%) | 92.1%<br>(77.5%-97.9%) | 94.7% |
| Zhengzhou Jinshui District General Hospital | 40 | 83.3%<br>(36.5%-99.1%) | 91.2%<br>(75.2%-97.7%) | 90.0% |
| Overall | 733 | 87.0%<br>(82.2%-90.7%) | 84.1%<br>(80.3%-87.2%) | 85.1% |

## Discussion

In this multi-center clinical study, 733 patients from five Chinese hospitals were involved in the statistical analysis. The overall OCT diagnosis results show a sensitivity of 87.0% (95% CI, 82.2%-90.7%), a specificity of 84.1% (95% CI, 80.3%-87.2%), and an accuracy of 85.1%. The highest sensitivity, specificity, and accuracy were all obtained at Luohe Central Hospital, which are 100.0%, 92.1%, and 94.7%, respectively. The lowest sensitivity (83.3%) was obtained at Zhengzhou Jinshui District General Hospital, and the lowest specificity (76.6%) and accuracy (81.1%) were both obtained at Liaoning Cancer Hospital & Institute. Since the pathology reports were issued by different hospitals, the diagnostic criteria may be slightly different among these hospitals, and this may be the main reason that caused the difference in the OCT diagnosis results. Moreover, the differences in sample size and patient population from the five hospitals may also lead to the variation of the OCT results. In general, this study shows that OCT can provide satisfactory diagnosis results which have good consistency with pathological results.

According to Zeng et al.’s study of ex-vivo OCM datasets and their corresponding histological sections, individual cells can be clearly identified in the OCM images, and five lesion categories are classified: normal cervix, ectropion, LSIL, HSIL, and invasive cervical lesions [17]. Compared with the OCM study by Zeng et al., OCT image features in this study are similar to those of OCM images. Cervical structural features of the five categories can also be well distinguished. All types of cervical tissues such as squamous epithelium, stroma, columnar epithelium, basal membrane, SCJ, cyst, gland, and tumor nest, can be observed, which are well-matched with the corresponding histological sections. With the high-resolution ability of OCT, low-risk and high-risk cervical lesions can be classified with high accuracy.

Some differences of image features between this OCT study and Zeng’s OCM study were found: (1) Fewer koilocytotic cell of LSIL tissues can be observed in OCT images, so the features of the LSIL tissues in most OCT images appear to be the same as those of normal tissues, but this does not affect the 2-class classification result. (2) The OCT images of some squamous cancer and some HSIL tissues share the icicle-like feature, which makes it hard to distinguish, but this does not affect the classification, either. (3) Since gland cancer usually occurs in the cervical canal which is inaccessible using the probe, adenocarcinoma tissues can be hardly observed in this in-vivo study.

Gallwas et al. showed that the sensitivity and specificity of OCT in the diagnosis of cervical intraepithelial lesions were 85%-98% and 39%-81%, respectively, and the high false-positive rate led to the decrease of specificity [20]. In the study by Zeng et al., a sensitivity of 80% and a specificity of 89% were obtained on ex-vivo specimens [17]. The sensitivity and specificity obtained in this in-vivo study are close to those reported by Zeng et al. This shows that OCT also works well on the in-vivo cervical diagnosis.

In this study, there are chances that the OCT results are inconsistent with corresponding pathological results. The possible reasons are as follows: (1) The scanning locations of OCT may not be exactly the same location for biopsy. The sampling differences may lead to mismatched results between OCT and histology. (2) Some HSIL diagnoses are based on focal lesions from the histological sections. These lesions may be missed in OCT images due to the sampling differences. (3) Some interference factors such as mucus, bubbles, and reflection worsen the quality of some OCT images, thus may affect the result of OCT classification.

As a potential cervical screening method, OCT was compared with the other two widely used methods: HPV test and TCT. It was reported that the HPV test has a high sensitivity of usually higher than 90% but a very low specificity of usually lower than 30% [21]. On the contrary, TCT can deliver a high specificity of usually higher than 85%, but a low sensitivity of usually lower than 60% [22]. In our study, it turns out that OCT has a slightly lower sensitivity but much higher specificity than the HPV test. Compared to TCT, OCT’s specificity is a bit lower, but its sensitivity is much higher. Among these three methods, OCT has the highest accuracy and the highest consistency with the gold standard. Besides, neither HPV nor TCT can provide real-time screening results or locate suspicious regions. While OCT can be used for real-time imaging. The results can be quickly acquired through image interpretation, and the location of lesions can be accurately located [23]. It needs to be noted that the recruited patients had either positive HPV results or suspicious cytology findings, so our OCT diagnosis results may not reflect the actual situation of the general population.

As a non-invasive real-time high-resolution in-vivo imaging technology, OCT has become more and more popular in clinical research. It has prospective clinical applications on in-vivo cervical examination. Meanwhile, OCT technology has certain limitations, such as the imaging depth. Moreover, rich experience is required to interpret OCT images. The classification training needs at least 4 weeks to achieve satisfactory accuracy. Ma’s study [24] showed that deep-learning-based CADx method outperformed human experts, and was also able to identify morphological characteristics in OCM images which were consistent with histopathological interpretations. Therefore, artificial intelligence (AI) aided diagnosis will be a great help in providing faster and more accurate OCT diagnosis.

Currently, OCT technology is still at an early stage in the study of cervical lesions. In the future, it needs to be improved in the following aspects: imaging resolution and quality, the ability of image interpretation, and introduction of AI aided diagnosis, as well as data acquisition on more clinical cases.

## Conclusion

In this multi-center clinical study on in-vivo cervical evaluation using a high-resolution OCT system, non-invasion and real-time examinations were performed with OCT. We found that OCT can not only provide high-resolution microscopic images with similar features to histology but also produce relatively precise diagnosis results compared to HPV test and TCT. It demonstrates the effectiveness of OCT technology in cervical lesion diagnosis. As a supplement of HPV and TCT, OCT has a promising potential to become a useful in-vivo cervical evaluation method.

## Data Availability

Some or all data used in the manuscript are available from the corresponding author by request.

## Abbreviations

3-D: 3-dimensional; AI: Artificial intelligence; ASCUS: atypical squamous cells of undetermined significance; CI: confidence interval; CIN: cervical intraepithelial neoplasia; H&E: hematoxylin and eosin; HPV: human papillomavirus; HSIL: high-grade squamous intraepithelial lesions; LSIL: low-grade squamous intraepithelial lesions; NC: Nabothian cysts; OCM: optical coherence microscopy; OCT: optical coherence tomography; SCJ: squamocolumnar junction; TCT: ThinPrep liquid-based cytology test.

## Acknowledgments

We thank all the doctors and nurses of the five hospitals for contribution in OCT examinations and diagnosis, and Ms. Qian Ma, Ms. Xing Dong, Ms. Xiaozhe Rong, Ms. Di Meng, Ms. Xian Cui, and Ms. Huan Xie for assistance in OCT image collecting, image processing, and comparison. This work is partially supported by the Henan Province medical science and technology research program co-sponsored by Henan province and the Ministry of Education (No. SBGJ2018050).

